# Increase of scabies infestations during the COVID-19 pandemic in Catalonia

**DOI:** 10.1101/2022.06.02.22275934

**Authors:** Manuel Medina, Núria Mora, Francisca Ramos, Leonardo Méndez-Boo, Carolina Guiriguet, Eduardo Hermosilla, Mireia Fàbregas, Ariadna Mas, Sara Rodoreda, Ermengol Coma, Francesc Fina

## Abstract

During the COVID-19 pandemic, several clinicians in Spain reported an increase in scabies diagnoses. We performed a time-series analysis with data from 2014 to 2022 to quantify this increase. We found an increasing trend during late 2020 and 2021, peaking in March 2022 with an almost 4.5-fold incidence than expected, especially in those aged between 16 and 30 years. Although scabies is more frequent in most socioeconomic deprived areas, the observed rise occurs in all the areas. We recommend increasing surveillance among other countries to detect unexpected increases in scabies outbreaks.

## Main text

Scabies, the contagious skin infestation caused by *Sarcoptes scabiei* that results in a severe itchy skin eruption, is a common disease worldwide and usually follows a seasonal pattern being more frequent in winter [1]. In recent years, some works have described an increase in scabies incidence during the first steps of the COVID-19 pandemic and before[1-6].Recently, several physicians reported a sudden rise in the cases of scabies in different Spanish regions that should be analyzed. We aim to quantify this increasing number of scabies infestations after two years of the beginning of the COVID-19 pandemic.

We extracted data on the number of scabies diagnoses from the primary care electronic health records (EHR) of the Catalan Institute of Health (ICS, its Catalan initials). ICS is the main primary care provider in Catalonia (Spain), and covers 85% of the primary care practices in the region, and about 6 million people. We included all scabies clinical diagnoses from January 2014 to April 2022 registered in the EHR according to the International Classification of Diseases 10th version (ICD-10) code B86. We performed a time series regression model adjusted by seasonality and trend using historical data from 2014 to 2018; and we obtained the expected incidence of scabies diagnoses with 95% confidence interval (95%CI). Time series was performed globally and by socioeconomic status (assessed using quartiles of the validated MEDEA indicator) [7]

Between January 2014 and April 2022, 85,153 scabies infestations were registered in the Catalan EHR. Of these, 48,556 (57%) have been reported since 2020. Monthly incidence ranged between 6 and 8 cases per 100,000 population during the 2014-2017 period; and then increased during the following years until it reached 52,8 cases per 100,000 in 2022. Figure 1 shows the trends during the whole study period by age groups. We observed an historical increasing trend, and a large growth which started in September 2020 and became more pronounced in September 2021, peaking in March 2022. The increase of scabies during the COVID-19 pandemic occurred across all age groups, although it was more relevant in those aged between 16 and 30 years. Figure 2 shows observed and expected monthly incidence of scabies since 2019. Observed scabies incidence was consistently above the estimate since the end of the lockdown in Spain (March-June 2020); and reached its peak in March 2022 with an incidence that was almost 4.5-fold higher than the expected. This led to an overall excess of diagnoses of 114.2% (95%CI: 82.1% - 159.9%) in 2021 and 281.7% (95%CI: 227.6% - 357.2%) in 2022 (Supplementary Table S1). Regarding socioeconomic status, although scabies is more frequent in most deprived areas, increasing trends were observed in all groups (Figure 2).

**Figure 1.**
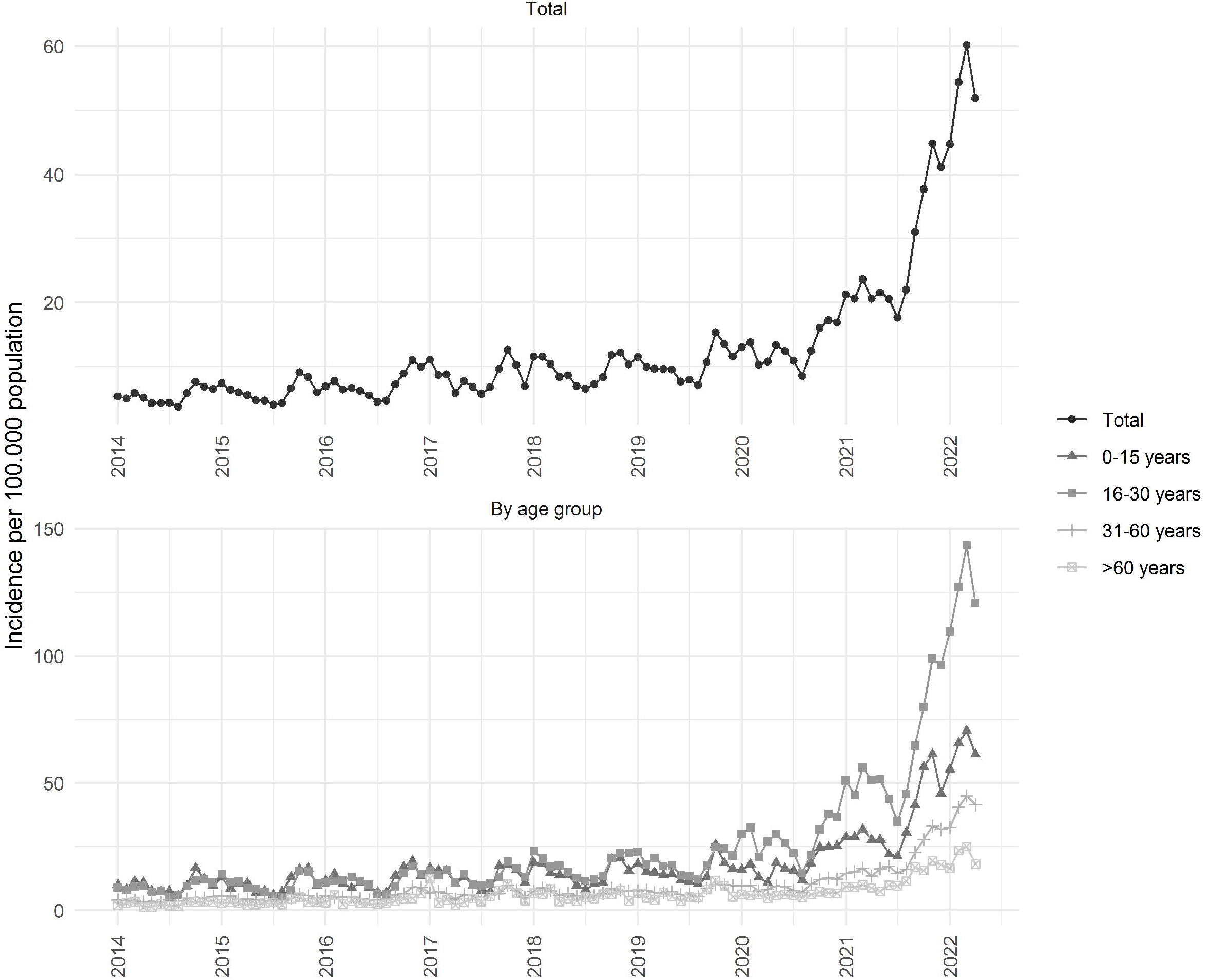
Monthly incidence of scabies infestations from January 2014 to April 2022 by age groups in Catalonia (Spain).

**Figure 2.**
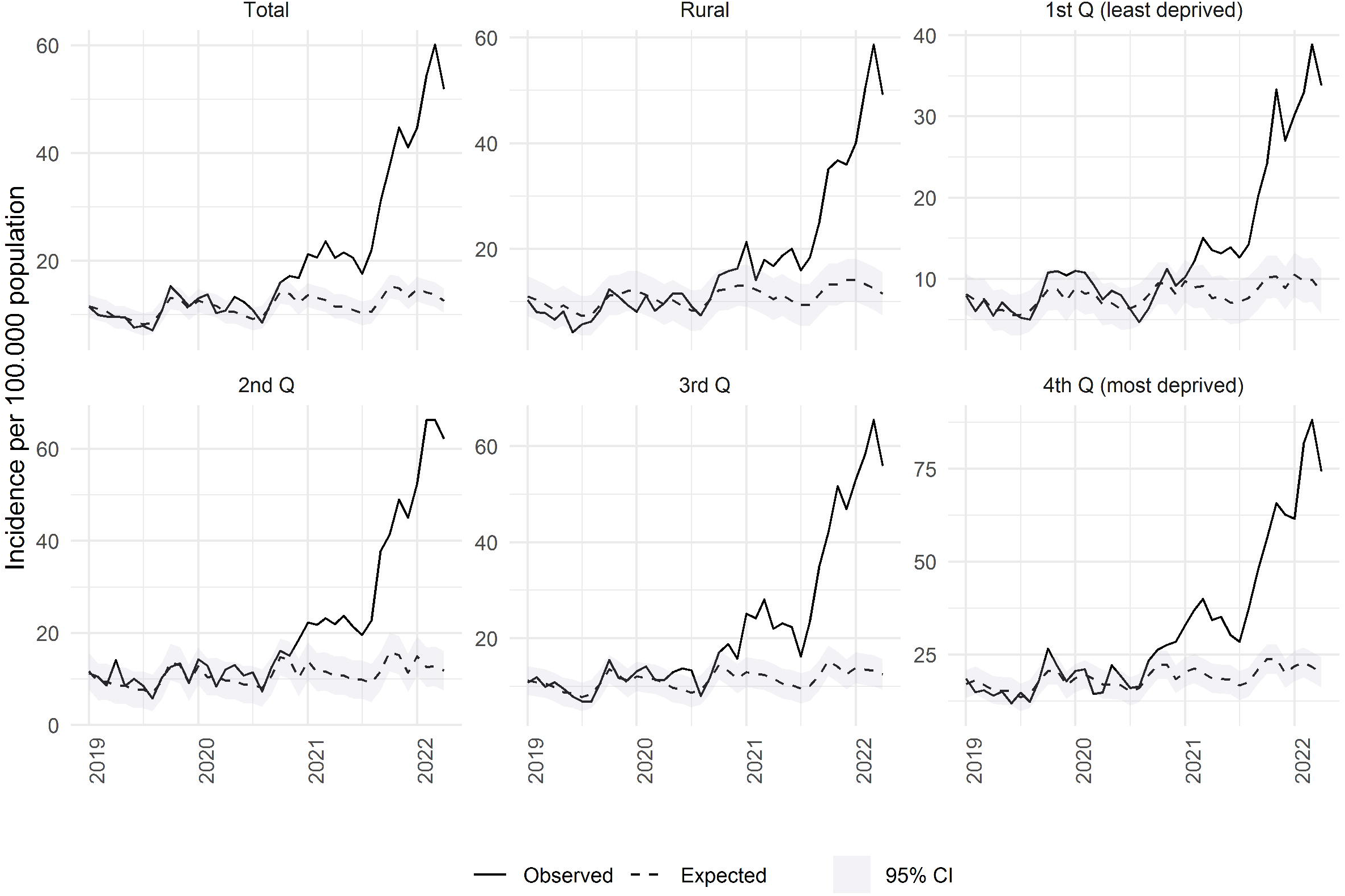
Observed and expected (with 95% CI) incidence of scabies infestations from January 2019 to April 2022 in Catalonia (Spain). Total and by rurality and socioeconomic status.

Other studies showed some increases during the first months of the COVID-19 pandemic but not to our extent [2-6]. In addition, although in a Spanish hospital the number of patients with scabies increased during the lockdown (March-June 2020) [3,6], we observed an expected number of diagnoses during those months that could be related to less primary care consultations as occurred with other relevant clinical conditions [8].

More studies are needed to assess which are the causes underlying the current increase of patients with scabies reported here, but several reasons could play a role. First, stay-at-home orders and quarantines probably contributed to more close contacts in households, increasing the risk of transmission [2,3,6,9]. Second, it has been suggested that daily habits including personal hygiene could have changed during the COVID-19 pandemic due to changes in social interactions, the increase of in-home working and the temporary closure of workplaces [2-9]. Not showering regularly and being unemployed were previously identified as risk factors for scabies infestations [2]. Third, in recent years, there has been concern about the possible permethrin resistance [10]. Four, it is possible that as a consequence of the impact of the pandemic on health care, many diagnoses were delayed and thus patients with scabies do not reach or delay the suitable treatment [4]. The detection and treatment of the index case is the most effective measure to stop the spread of the parasite [3,4,6]. Five, there could exist some reasons linked to changes in health-seeking behavior of patients due to the fear of being infected by SARS-CoV-2 [8]. Finally, some healthcare professionals suspected that the loss of treatment efficacy would be attributable to treatment noncompliance and treatment failures, rather than permethrin resistance [3,10,11].

Limitations of this analysis include the use of EHR data and alterations in patients’ healthcare-seeking behavior during COVID-19 pandemic that could change the distribution of the outpatients.

Our results should be taken as a warning signal for other countries to increase scabies surveillance to ensure outbreaks are under control.

## Data Availability

EHR data and analytical code are provided at https://github.com/ErmengolComa/sarna/

## Statements

### Ethics approval

This study is part of a project that analyses the impact of COVID-19 pandemic, related control measures and organizational changes on primary care. The project was approved by the Clinical Research Ethics Committee of the IDIAP Jordi Gol (project code 20/172-PCV).

### Data availability statement

EHR data and analytical code are provided at https://github.com/ErmengolComa/sarna/

### Author’s contributions

EC, CG, NM, MM, LM-B, FR, MF, FF, AM and SR contributed to the design of the study, interpretation of the results and reviewed the manuscript. EC, FF and NM had access to the data, performed the statistical analysis and acted as guarantors. EC, CG, MM, LM-B, FR and NM wrote the first draft of the manuscript. All authors critically revised the manuscript. The corresponding author attests that all listed authors meet the authorship criteria and that no others meeting the criteria have been omitted.

