## Supplementary material for "Increase of scabies infestations during the COVID-19 pandemic in Catalonia"

**Supplementary Table S1. Percentage of excess (with 95%CI) of scabies diagnoses according to the expected since 2019 toin Catalonia (Spain)**

|  | 2019 | 2020 | 2021 | 2022 |
| --- | --- | --- | --- | --- |
| **Total** | **NE** | **NE** | **114.2 (82.1 - 159.9)** | **281.7 (227.6 - 357.2)** |
| **Sex** | | | | |
| Women | NE | NE | 116.6 (79.3 - 173.5) | 288.5 (224.6 - 383.6) |
| Men | NE | NE | 111.6 (79.5 - 157.5) | 274.8 (221.5 - 349.2) |
| **Age groups** | | | | |
| 0 -15 years | NE | NE | 94.3 (56.7 - 155.6) | 215.7 (157.8 - 307.1) |
| 16 -30 years | NE | 35.1 (12 - 70.1) | 166.3 (123.3 - 229.8) | 390.4 (317.2 - 494.9) |
| 31 -60 years | NE | NE | 112.4 (75.1 - 169.9) | 294.1 (227.2 - 395.5) |
| >60 years | NE | NE | NE | 124.7 (57.3 - 293.2) |
| **Socioeconomic status** | | | | |
| First quartile: urban least  deprived | NE | NE | 102.8 (55.2 - 192.3) | 251.4 (174 - 389.5) |
| Second  quartile | NE | NE | 146.6 (82.4 - 280.6) | 373.2 (256.1 - 605.2) |
| Third  quartile | NE | NE | 149.8 (98.1 - 238) | 338.5 (252.5 - 480) |
| Fourth  quartile: urban more deprived | NE | NE | 111.9 (77.1 - 163.8) | 253.3 (197.6 - 334.7) |
| Rural areas | NE | NE | 97.8 (47.5 - 200.3) | 285.4 (192.2 - 465.9) |

NE: No excess
